# Tertiary-care Management of TB/HIV Co-infection in Port Harcourt, Nigeria: A Retrospective Cohort Analysis

**DOI:** 10.1101/2025.05.22.25328150

**Authors:** Prince C. Nnadozie, Sylva C. Nnadozie, Abimbola T. Awopeju, Inumanye N. Ojule

## Abstract

**Background:** Monitoring the treatment of tuberculosis(TB) and human immunodeficiency virus(HIV) co-infection is very important to improve patient management and compliance. This study assessed the treatment outcomes of registered TB/HIV co-infected patients enrolled in the Directly Observed Treatment Short course(DOTS) program.

**Methods:** A retrospective study was conducted in a tertiary-care hospital in Port Harcourt, Nigeria from January 1, 2014 to December 31, 2019. Data were extracted from the National TB program register and analyzed using the statistical software package(IBM SPSS version30.0). Bivariate and multivariate logistic regressions were used to identify factors associated with treatment outcomes at 95% confidence intervals.

**Results:** Among 305(37.4%) TB/HIV co-infected patients, the mean age was 29.2years, with 29.8% in the 0-14 age group, 89.5% new cases, and 89.2% pulmonary TB(PTB) cases. Males(50.2%) slightly outnumbered females(49.8%). Antiretroviral therapy(ART) and cotrimoxazole preventive therapy(CPT) uptake were high(95.1% and 93.4%, respectively). Treatment outcomes were: 19.0% cured, 23.3% completed treatment, 0.3% failed treatment, 23.6% died, 14.8% lost to follow-up, 10.8% transferred out, and 8.2% not evaluated. Overall, treatment success rate was 52.2%. Age(15-24 years: AOR=6.35, 95%CI:1.56-25.86; 25-34 years: AOR=3.97, 95%CI:1.37-11.45; and 45-54 years: AOR=6.40, 95%CI:2.27-18.06) and alternative diagnostic method(AOR=0.32, 95%CI:0.12-0.76) were significantly associated with unsuccessful treatment outcomes.

**Conclusion:** The treatment success rate of TB/HIV co-infected patients was relatively low and fell below the WHO target despite high uptake of ART and CPT. Study underscores the need for integrating targeted interventions, including alternative diagnostic methods, into clinical practice and policy to improve treatment outcomes among TB/HIV co-infected patients, particularly in specific age groups.

## Introduction

Nigeria’s tertiary-care facilities are fighting a formidable battle against tuberculosis (TB) and human immunodeficiency virus (HIV) infections. Despite ongoing efforts to manage these two major interconnected public health challenges, Nigeria’s situation remains concerning. Globally, progress in combating TB and HIV infections is marked by tremendous setbacks and retrogression. According to the World Health Organization’s (WHO) *Global Tuberculosis Report* 2023, 10.6 million people worldwide contracted TB in 2022, with 6.3% of those cases involving people living with HIV (PLWHIV) [1]. This indicates an increase from the estimated 10 million global TB cases in 2019, of which 8.2% involved PLWHIV [2].

Multiple factors contribute to the rising prevalence of TB/HIV co-infection, its morbidity and mortality. Among these include the COVID-19 pandemic, poverty, drug resistance, and stigmatization [3-6]. TB and HIV are closely linked, causing deleterious effects in co-infected patients [7]. If left untreated, these infections compromise the immune system and reduce patients’ lifespan and quality of life [8, 9]. While TB can influence disease progression in HIV positive patients, HIV infection can cause TB progression from the latent phase to the active phase and increase the incidence of TB[10,11].

Effective management of TB/HIV co-infection is crucial in the monitoring of treatment outcomes [12,13]. However, data on treatment outcomes for TB/HIV co-infections in Nigeria is limited [14]. While some studies reported successful treatment outcomes in tertiary health facilities in Nigeria [15,16], others revealed poor outcomes [17-20], affirming the potential for TB/HIV co-infection to undermine public health gains. Hence, this study evaluated treatment outcomes for TB/HIV co-infected patients registered in a tertiary-care facility in Port Harcourt, Nigeria. Understanding the epidemiology of TB/HIV co-infection in Nigeria can inform policy decisions, enable efficient resource allocation, enhance treatment outcomes, reduce illness and death rates, and prevent the spread of drug-resistant TB strains.

## Methods

### Study design, setting and target population

This study employed a retrospective, descriptive and analytic design to assess treatment outcomes of all registered TB/HIV co-infected patients enrolled at University of Port Harcourt Teaching Hospital (UPTH), from 1st January 2014 to 31st December 2019. This tertiary care hospital is located behind the University of Port Harcourt, Port Harcourt, Rivers State, Nigeria. It is one of the biggest federal government of Nigeria referral hospitals in Niger Delta region of Southern Nigeria, consisting of a HIV/AIDS clinic, and a TB treatment facility that is set up in accordance with the National Tuberculosis and Leprosy Control Program (NTBLCP) [21]. In addition to several private healthcare centers in Port Harcourt, there were other government referral hospitals in Port Harcourt belonging to Rivers State Government which also have Directly Observed Treatment Short-course (DOTS) facilities for the treatment of TB patients. Sometimes patients are being referred from these other hospitals to UPTH for treatment. Port Harcourt is an urban metropolis, mostly dominated by Christians [22], with an estimated population of 3,020,000 in 2020 (5.12% increase from 2019) [23,24]. It is comprised mostly of a heterogenous group of people of different tribes and ethnicity, majority are the coastal people including Ikwerres, Ijaws, Ogonis, and among others [25]. Many people who live in Port Harcourt are educated and speak English [25]. People living in rural areas sometimes visit UPTH or these other referral centers in Port Harcourt for healthcare services. The study area is in an oil rich city with multinational oil companies whose business activities result in gas flaring and other sundry environmental pollution.

### Eligibility criteria and study variables

TB/HIV co-infected patients of all age groups (in years) who had started and completed a course of anti-TB treatment, or incomplete medical records (treatment outcome unknown), or transferred out during the study period were included in the study. Patients with multidrug-resistant/extensively drug-resistant tuberculosis (MDR/XDR-TB) were excluded from the study because they required specialized treatment regimens that are different from those used for drug-susceptible TB. Also, this study excluded patients transferred into UPTH from other health facilities with no baseline data. The outcome variable was treatment outcome, and could only one of the following: cured, treatment completed, treatment failure, died, loss to follow-up (defaulter), transferred out, or not evaluated. The determinant factors that we had included: age (years), sex, HIV patients receiving care/support (antiretroviral therapy, ART, and cotrimoxazole preventive therapy, CPT), TB treatment regimen (standard: 2 months intensive phase of treatment with HRZE and continuation of treatment with HR for an additional 4 months; retreatment cases: 2 months initial treatment with SHRZE, and followed by 1 month treatment with HRZE and 3 months completion of treatment with HRE)(S: streptomycin, H:isoniazid, R:rifampicin, Z:pyrazinamide, E:ethambutol), diagnostic methods used for TB investigation (microscopy, Xpert MTB/RIF, culture, and other alternatives), type of TB (PTB, and EPTB), and type of patient (new, and retreatment). Socioeconomic status, education, employment, and history about comorbidities were not available in the registers.

### Data collection, management and statistical analysis

We accessed patients data on 16.02.2021 and conducted a review on 816 TB patients who had registered on national TB program (NTP) at UPTH between January 1, 2014, to December 31, 2019. Data of all new and retreatment TB/HIV co-infected patients were extracted from the treatment records. Relevant information about age, sex, type of TB patients, form of TB, diagnostic method used, HIV patients on ART and CPT, treatment regimen used, and treatment outcomes were collected and entered using EpiData software (version 4.6.02). The data for patient’s age were categorized following standard WHO age groups for TB/HIV reports, and was as follows: 0 – 14, 15 – 24, 25 – 34, 35 – 44, 45 – 54, and ≥55 years. As part of the data management procedure, cleaning, coding and recoding of extracted data were carried out by two researchers. Error entries were corrected where possible to ensure the consistency and accuracy of data. The study utilized descriptive statistics to summarize the clinical and demographic characteristics of the study population. Test of association between the independent variables and the main outcomes of the study was conducted using the Chi-square (χ2) test. For variables with an expected count of less than five, Fisher’s exact test was reported [12,26]. To identify demographic and clinical characteristics variables associated with TB/HIV treatment outcomes, a multivariate logistic regression analysis was run. The treatment outcome variables “*cured, completed treatment, failed treatment, died, lost to follow-up, transferred out, and not evaluated”* were recoded into a dichotomic variable: 0 = successful outcome (cured + completed treatment) and 1 = unsuccessful outcome (failed treatment + died + lost to follow-up). We excluded *“transferred out”* and *“not evaluated”* cases during the analysis stage. Not evaluated cases may not have been assessed for treatment outcome and including them in the analysis could bias the results. Whereas patients who were transferred to another facility or program may not have completed their treatment and may not necessarily be a treatment failure. Therefore, including them could overestimate the number of unsuccessful outcomes. The effect measure for the outcome was Odds ratio. Crude and adjusted odds ratios (COR and AOR, respectively) were computed and used to see the strength of association. All analyses were performed at 5% level of significance using IBM statistical package for social sciences (SPSS) version 30.0 IBM SPSS (Armonk, NY, USA).

### Ethical considerations

Ethical approval (with reference number UPTH/ADM/90/S.II/Vol.XI/1067) for this study was approved by UPTH Ethics and Research Committee (ERC) in Nigeria, with a recommendation by Regional Committee for Medical and Health Research Ethics (REK) in Norway (reference number: 154120). Patients’ consent, before reviewing records, was not obtained or necessary for this study and usually not recommended by the Ethics Committee.

### Operational definitions

a. **Treatment outcome:** The components of treatment outcomes were defined according to WHO [27-29].
  i. **Cured:** A registered patient who was smear or culture positive at diagnosis, but on treatment completion, shows smear or culture negative in the last month of treatment and at least one previous occasion during treatment.
  ii. **Treatment completed:** A registered TB patient who completed treatment without bacteriological evidence of failure or cure, either because tests were not done or because results were unavailable.
  iii. **Treatment failure:** A registered TB patient who remained sputum smear or culture positive at 5th month or later during treatment.
  iv. **Died:** A registered TB patient who died for any reason before starting or during the course of treatment.
  v. **Loss to follow-up (defaulter):** A registered TB patient whose treatment was interrupted for two consecutive months or more.
  vi. **Transferred out:** A registered TB patient who has been transferred to another treatment center or hospital, but whose definitive outcome at the end of treatment was not established.
  vii. **Not evaluated:** Refers to a registered patient with missing information who no treatment outcome was assigned. This includes cases for whom the treatment outcome were unknown to the reporting unit.
b. **Transferred in:** A registered patient who has been transferred from another unit or hospital and whose baseline data were not available in the TB register.
c. **Treatment success (successful outcome):** This is defined as the total number of all registered patients that were cured and all those whose TB treatment were completed (i.e., cured + treatment completed).
d. **Types of patients:** This refers to new, and retreatment (with relapse, return after lost to follow-up, return after failure, others) cases.
  i. **New case:** A registered TB patient without or with less than one month of previous treatment [30,31].
  ii. **Retreatment case:** According to WHO definition [30], this includes:
  - *Relapse patient:* A bacteriologically confirmed positive registered TB patient who was previously treated and declared cured, or treatment completed at the end of the most recent course of anti-TB treatment and was at the present time diagnosed with a recurrent episode of TB (either a true relapse or a new episode of TB caused by reinfection).
  - *Treatment after failure patient:* A bacteriologically confirmed positive registered TB patient who had previously been treated for TB and whose treatment failed at the end of their most recent course of treatment.
  - *Treatment after loss to follow-up (default) patient:* A bacteriologically confirmed positive registered TB patient who had previously been treated for TB and was declared lost to follow-up at the end of their most recent course of treatment.
  - *Other previously treated patient:* A patient who had previously been treated for TB, and get TB again, but are not sputum confirmed.
e. **Unsuccessful treatment outcome:** This is defined as the sum of all registered patients who were not successfully treated (i.e., failed treatment + dead + lost to follow-up).

## Results

### Demographic characteristics of TB/HIV co-infected patients

Of the 816 TB patients that were screened for HIV infection within the study period, a total of 305 (37.4%) had HIV infection. Table 1 shows the demographic and clinical characteristics of the study participants. The mean age (± standard deviation, range) of participants was 29.2(±18.9, 1 – 72) years and median age was 34 years. The age group 0-14 years exhibited the highest prevalence (29.8%), closely followed by the 35-44 years age group (27.2%). In contrast, the 15-24 years age group had the least prevalence rate (5.2%). The year 2015 recorded the highest TB/HIV prevalence (23.3%), subsequently followed by 2014 (19.3%), 2016 (16.4%), 2017/2019 (14.1%), and 2018 (12.8%). Notably, males (50.2%) slightly outnumbered females (49.8%) among the TB/HIV co-infected patients.

**Table 1:** Demographic and clinical characteristics of TB/HIV co-infected patients in UPTH, Nigeria between 2014 and 2019, stratified by year (N=305)

| Variable | 2014<br>n(%) | 2015<br>n(%) | 2016<br>n(%) | 2017<br>n(%) | 2018<br>n(%) | 2019<br>n(%) | All patients<br>n(%) |
| --- | --- | --- | --- | --- | --- | --- | --- |
| **Age (years)** |  |  |  |  |  |  |  |
| 0 - 14 | 23(39.0) | 21(29.6) | 17(34.0) | 12(27.9) | 6(15.4) | 12(27.9) | 91(29.8) |
| 15 - 24 | 2(3.4) | 5(7.0) | 4(8.0) | 1(2.3) | 1(2.6) | 3(7.0) | 16(5.2) |
| 25 - 34 | 7(11.9) | 15(21.1) | 6(12.0) | 4(9.3) | 7(17.9) | 8(18.6) | 47(15.4) |
| 35 - 44 | 14(23.7) | 13(18.3) | 16(32.0) | 11(25.6) | 17(43.6) | 12(27.9) | 83(27.2) |
| 45 - 54 | 10(16.9) | 12(16.9) | 3(6.0) | 10(23.3) | 7(17.9) | 6(14.0) | 48(15.7) |
| ≥55 | 3(5.1) | 5(7.0) | 4(8.0) | 5(11.6) | 1(2.6) | 2(4.7) | 20(6.6) |
| **Total** | **59(19.3)** | **71(23.3)** | **50(16.4)** | **43(14.1)** | **39(12.8)** | **43(14.1)** | **305(100.0)** |
| **Sex** |  |  |  |  |  |  |  |
| Male | 23(39.0) | 36(50.7) | 26(52.0) | 23(53.5) | 24(61.5) | 21(48.8) | 153(50.2) |
| Female | 36(61.0) | 35(49.3) | 24(48.0) | 20(46.5) | 15(38.5) | 22(51.2) | 152(49.8) |
| **Total** | **59(19.3)** | **71(23.3)** | **50(16.4)** | **43(14.1)** | **39(12.8)** | **43(14.1)** | **305(100.0)** |
| **Type of TB** |  |  |  |  |  |  |  |
| PTB | 52(88.1) | 57(80.3) | 49(98.0) | 39(90.7) | 34(87.2) | 41(95.3) | 272(89.2) |
| EPTB | 7(11.9) | 7(9.9) | 1(2.0) | 4(9.3) | 5(12.8) | 2(4.7) | 26(8.5) |
| Missing | 0(0.0) | 7(9.9) | 0(0.0) | 0(0.0) | 0(0.0) | 0(0.0) | 7(2.3) |
| **Total** | **59(19.3)** | **71(23.3)** | **50(16.4)** | **43(14.1)** | **39(12.8)** | **43(14.1)** | **305(100.0)** |
| **Type of patient** |  |  |  |  |  |  |  |
| New | 52(88.1) | 64(90.1) | 42(84.0) | 37(86.0) | 36(92.3) | 42(97.7) | 273(89.5) |
| Retreatment relapse | 1(1.7) | 2(2.8) | 3(6.0) | 5(11.6) | 3(7.7) | 0(0.0) | 14(4.6) |
| Retreatment after failure | 2(3.4) | 0(0.0) | 0(0.0) | 0(0.0) | 0(0.0) | 0(0.0) | 2(0.7) |
| Retreatment after loss to follow-up | 2(3.4) | 2(2.8) | 2(4.0) | 1(2.3) | 0(0.0) | 0(0.0) | 7(2.3) |
| Retreatment others | 2(3.4) | 3(4.2) | 2(4.0) | 0(0.0) | 0(0.0) | 0(0.0) | 7(2.3) |
| Missing | 0(0.0) | 0(0.0) | 1(2.0) | 0(0.0) | 0(0.0) | 1(2.3) | 2(0.7) |
| **Total** | **59(19.3)** | **71(23.3)** | **50(16.4)** | **43(14.1)** | **39(12.8)** | **43(14.1)** | **305(100.0)** |
| **HIV care & support** |  |  |  |  |  |  |  |
| Not on ART | 2(3.4) | 0(0.0) | 0(0.0) | 4(9.3) | 2(5.1) | 7(16.3) | 15(4.9) |
| On ART | 57(96.6) | 71(100.0) | 50(100.0) | 39(90.7) | 37(94.9) | 36(83.7) | 290(95.1) |
| **Total** | **59(19.3)** | **71(23.3)** | **50(16.4)** | **43(14.1)** | **39(12.8)** | **43(14.1)** | **305(100.0)** |
| Not on CPT | 6(10.2) | 1(1.4) | 0(0.0) | 4(9.3) | 2(5.1) | 7(16.3) | 20(6.6) |
| On CPT | 53(89.8) | 70(98.6) | 50(100.0) | 39(90.7) | 37(94.9) | 36(83.7) | 285(93.4) |
| **Total** | **59(19.3)** | **71(23.3)** | **50(16.4)** | **43(14.1)** | **39(12.8)** | **43(14.1)** | **305(100.0)** |
| **Regimen** |  |  |  |  |  |  |  |
| Standard (2HRZE/4HR) | 44(74.6) | 67(94.4) | 50(100.0) | 42(97.7) | 38(97.4) | 43(100.0) | 284(93.1) |
| Retreatment(2SHRZE/1HRZE/3HRE) | 15(25.4) | 4(5.6) | 0(0.0) | 1(2.3) | 1(2.6) | 0(0.0) | 21(6.9) |
| **Total** | **59(19.3)** | **71(23.3)** | **50(16.4)** | **43(14.1)** | **39(12.8)** | **43(14.1)** | **305(100.0)** |
| **Method of Diagnosis** |  |  |  |  |  |  |  |
| Smear microscopy | 32(54.2) | 37(52.1) | 22(44.0) | 10(23.3) | 2(5.1) | 1(2.3) | 104(34.1) |
| Xpert MTB/RIF | 0(0.0) | 2(2.8) | 10(20.0) | 13(30.2) | 20(51.3) | 23(53.5) | 68(22.3) |
| Alternatives | 26(44.1) | 31(43.7) | 15(30.0) | 19(44.2) | 14(35.9) | 19(44.2) | 124(40.7) |
| Missing | 1(1.7) | 1(1.4) | 3(6.0) | 1(2.3) | 3(7.7) | 0(0.0) | 9(3.0) |
| **Total** | **59(19.3)** | **71(23.3)** | **50(16.4)** | **43(14.1)** | **39(12.8)** | **43(14.1)** | **305(100.0)** |
| **Treatment outcome of patients** |  |  |  |  |  |  |  |
| Cured | 6(10.2) | 13(18.3) | 11(22.0) | 12(27.9) | 7(17.9) | 9(20.9) | 58(19.0) |
| Completed treatment | 22(37.3) | 17(23.9) | 11(22.0) | 7(16.3) | 11(28.2) | 3(7.0) | 71(23.3) |
| Failed treatment | 1(1.7) | 0(0.0) | 0(0.0) | 0(0.0) | 0(0.0) | 0(0.0) | 1(0.3) |
| Died | 7(11.9) | 20(28.2) | 14(28.0) | 13(30.2) | 8(20.5) | 10(23.3) | 72(23.6) |
| Lost to follow-up | 18(30.5) | 5(7.0) | 9(18.0) | 4(9.3) | 4(10.3) | 5(11.6) | 45(14.8) |
| Transferred out | 5(8.5) | 11(15.5) | 4(8.0) | 6(14.0) | 4(10.3) | 3(7.0) | 33(10.8) |
| Not evaluated | 0(0.0) | 5(7.0) | 1(2.0) | 1(2.3) | 5(12.8) | 13(30.2) | 25(8.2) |
| **Total** | **59(19.3)** | **71(23.3)** | **50(16.4)** | **43(14.1)** | **39(12.8)** | **43(14.1)** | **305(100.0)** |

TB: tuberculosis; HIV: human immunodeficiency virus; PTB: pulmonary TB; EPTB: extrapulmonary TB; ART: antiretroviral therapy; CPT: cotrimoxazole preventive therapy; regimen 1: 2HRZE/4HR, regimen 2: 2SHRZE/1HRZE/3HRE (1, 2, 3, 4: months, R: rifampicin, H: isoniazid, Z: pyrazinamide, Ethambutol; and S: streptomycin); MTB: Mycobacterium TB; RIF: resistance to rifampicin; and Alternatives: clinical diagnosis, score charts, chest X-Ray, magnetic resonance imaging (MRI), Mantoux test, or tissue pathology; N: total number of participants in the study; n: frequency; %: percentage of frequency.

### Clinical characteristics

PTB was the predominant form of TB, affecting 89.2% of patients, while 8.5% had EPTB (Table 1). The majority of patients (89.5%) were new cases, whereas 9.8% were retreatment cases. A significant proportion of patients received ART (95.1%) and CPT (93.4%). Furthermore, 93.1% of patients completed the standard 6-month TB treatment regimen (2HRZE/4HR), while 6.9% underwent retreatment regimens (2SHRZE/1HRZE/3HRE). Diagnostic approaches varied, with 40.7% of patients diagnosed through: clinical examination, score charts, chest X-Ray, magnetic resonance imaging (MRI), Mantoux test, or tissue pathology. Xpert MTB/RIF, and smear microscopy for acid fast bacilli was used in the diagnosis of 22.3% and 34.1% of TB/HIV co-infected patients, respectively.

### Treatment outcomes

The treatment outcomes of TB/HIV co-infected patients were as follows: 58 (19.0%) were cured, 71 (23.3%) completed treatment, 1 (0.3%) failed treatment, 72 (23.6%) died, 45 (14.8%) were lost to follow-up, 33 (10.8%) were transferred out, and 25 (8.2%) were not evaluated (Table 1). The study observed an overall treatment success rate of 52.2% (129/247) and an unsuccessful treatment outcome rate of 47.8% (118/247). The bivariate analysis revealed that HIV care and support, specifically being on ART (χ2 = 8.56, p=0.003) and CPT (χ2 = 6.65, p=0.01), were significantly associated with treatment outcomes (Table 2). Annually, the treatment success rates ranged from 44.4% (2019) to 60.0% (2018) (χ2 = 1.71, p=0.89). Patients who received ART and CPT achieved nearly equal rates of successful treatment outcomes (approximately 54%). However, patients who did not receive ART or CPT substantially had a higher rate of unsuccessful outcomes (90.9% and 80%), respectively. Unsuccessful treatment outcomes were more common among TB/HIV co-infected patients who were male (53.7%), diagnosed through alternative ways (58.1%), and those who fell under the age bracket of 45-54 years (64.3%).

**Table 2:** Demographic and clinical characteristics of TB/HIV co-infected patients in UPTH, Nigeria between 2014 and 2019, stratified by TB treatment outcomes (N=247)

| Variable | Treatment outcome |  | p-Value |
| --- | --- | --- | --- |
|  | Successful outcome<br>n(%) | Unsuccessful outcome<br>n(%) |  |
| **Age (years)** |  |  |  |
| 0 - 14 | 43(60.6) | 28(39.4) | 0.09 |
| 15 - 24 | 5(38.5) | 8(61.5) |  |
| 25 – 34 | 20(52.6) | 18(47.4) |  |
| 35 – 44 | 39(59.1) | 27(40.9) |  |
| 45 – 54 | 15(35.7) | 27(64.3) |  |
| ≥55 | 7(41.2) | 10(58.8) |  |
| **Sex** |  |  |  |
| Male | 57(46.3) | 66(53.7) | 0.07 |
| Female | 72(58.1) | 52(41.9) |  |
| **Type of TB** |  |  |  |
| PTB | 119(53.1) | 105(46.9) | 0.83 |
| EPTB | 10(43.5) | 13(56.5) |  |
| **Type of patient** |  |  |  |
| New | 114(52.1) | 105(47.9) | 0.59 |
| Retreatment | 15(53.6) | 13(46.4) |  |
| **HIV care & support** |  |  |  |
| Not on ART | 1(9.1) | 10(90.9) | 0.003* |
| On ART | 128(54.2) | 108(45.8) |  |
| Not on CPT | 3(20.0) | 12(80.0) | 0.01* |
| On CPT | 126(54.3) | 106(45.7) |  |
| **Regimen** |  |  |  |
| Standard (2HRZE/4HR) | 120(52.9) | 107(47.1) | 0.50 |
| Retreatment(2SHRZE/1HRZE/3HRE) | 9(45.0) | 11(55.0) |  |
| **Method of Diagnosis** |  |  |  |
| Smear microscopy | 57(62.0) | 35(38.0) | 0.49 |
| Xpert MTB/RIF | 28(56.0) | 22(44.0) |  |
| Alternatives | 44(41.9) | 61(58.1) |  |
| **Year of diagnosis/treatment** |  |  |  |
| 2014 | 28(51.9) | 26(48.1) | 0.89 |
| 2015 | 30(54.5) | 25(45.5) |  |
| 2016 | 22(48.9) | 23(51.1) |  |
| 2017 | 19(52.8) | 17(47.2) |  |
| 2018 | 18(60.0) | 12(40.0) |  |
| 2019 | 12(44.4) | 15(55.6) |  |

TB: tuberculosis; HIV: human immunodeficiency virus; PTB: pulmonary TB; EPTB: extrapulmonary TB; ART: antiretroviral therapy; CPT: cotrimoxazole preventive therapy; regimen 1: 2HRZE/4HR, regimen 2: 2SHRZE/1HRZE/3HRE (1, 2,3, 4: months, R: rifampicin, H: isoniazid, Z: pyrazinamide, Ethambutol; and S: streptomycin); MTB: Mycobacterium TB; RIF: resistance to rifampicin; and Alternatives: clinical diagnosis, score charts, chest X-Ray, magnetic resonance imaging (MRI), Mantoux test, or tissue pathology; N: total number of participants in the study; n: frequency; %: percentage of frequency; Asterisks (\*):statistically significant at $p \leq 0.05$ .

### Regression analysis

Multivariate logistic regression analyses were conducted to assess the relationship between unsuccessful treatment outcomes, and demographic and clinical characteristics among TB/HIV co-infected patients (Table 3). The odds of unsuccessful treatment outcomes were evaluated across different years, with 2014 as the reference year. Patients treated in 2015 had slightly lower odds of unsuccessful treatment outcomes (COR = 0.90, 95%CI: 0.42-1.91; AOR = 0.94, 95%CI: 0.39-2.28). A higher likelihood of unsuccessful treatment outcome was observed in patients treated in 2016 (COR = 1.13, 95%CI: 0.51-2.49; AOR = 1.30, 95%CI: 0.51-3.35). The likelihood of unsuccessful treatment outcomes decreased in 2017 (COR = 0.96, 95%CI: 0.41-2.24; AOR = 0.63, 95%CI: 0.22-1.82). A further decrease in the odds of unsuccessful treatment outcome was observed in 2018 (COR = 0.72, 95%CI: 0.29-1.77; AOR = 0.49, 95%CI: 0.15-1.60). Although the COR suggested an increased likelihood of unsuccessful outcome (1.35, 95%CI: 0.53-3.41), the AOR indicated a 31% lower odds of unsuccessful treatment outcomes in 2019 (0.69, 95%CI: 0.20-2.37).

**Table 3:** Predictors of unsuccessful treatment outcomes among TB/HIV co-infected patients in UPTH, Nigeria between 2014 and 2019.

| Variable | Crude OR (95% CI) | Adjusted OR (95% CI) |
| --- | --- | --- |
| **Age (years)** |  |  |
| 0 - 14 | 1.00 | 1.00 |
| 15 - 24 | 2.46(0.73-8.28) | 6.35(1.56-25.86) |
| 25 – 34 | 1.38(0.62-3.06) | 3.97(1.37-11.45) |
| 35 – 44 | 1.06(0.54-2.11) | 2.52(0.98-6.48) |
| 45 – 54 | 2.76(1.25-6.09) | 6.40(2.27-18.06) |
| ≥55 | 2.19(0.75-6.44) | 3.65(0.90-14.80) |
| **Sex** |  |  |
| Male | 1.00 | 1.00 |
| Female | 0.62(0.38-1.03) | 0.71(0.40-1.27) |
| **Type of TB** |  |  |
| PTB | 1.00 | 1.00 |
| EPTB | 1.11(0.44-2.76) | 0.67(0.21-2.12) |
| **Type of patient** |  |  |
| New | 1.00 | 1.00 |
| Retreatment | 0.80(0.35-1.81) | 0.62(0.23-1.72) |
| **HIV care & support** |  |  |
| Not on ART | 1.00 | 1.00 |
| On ART | 0.08(0.01-0.67) | 0.11(0.01-1.51) |
| Not on CPT | 1.00 | 1.00 |
| On CPT | 0.21(0.06-0.77) | 0.79(0.13-4.82) |
| **Regimen** |  |  |
| Standard (2HRZE/4HR) | 1.00 | 1.00 |
| Retreatment(2SHRZE/1HRZE/3HRE) | 1.37(0.55-3.44) | 1.75(0.47-6.54) |
| **Method of Diagnosis** |  |  |
| Smear microscopy | 1.00 | 1.00 |
| Xpert MTB/RIF | 1.28(0.64-2.58) | 1.65(0.64-4.27) |
| Alternatives | 0.73(0.19-0.94) | 0.32(0.12-0.76) |
| **Year of diagnosis/treatment** |  |  |
| 2014 | 1.00 | 1.00 |
| 2015 | 0.90(0.42-1.91) | 0.94(0.39-2.28) |
| 2016 | 1.13(0.51-2.49) | 1.30(0.51-3.35) |
| 2017 | 0.96(0.41-2.24) | 0.63(0.22-1.82) |
| 2018 | 0.72(0.29-1.77) | 0.49(0.15-1.60) |
| 2019 | 1.35(0.53-3.41) | 0.69(0.20-2.37) |

TB: tuberculosis; HIV: human immunodeficiency virus; PTB: pulmonary TB; EPTB: extrapulmonary TB; ART: antiretroviral therapy; CPT: cotrimoxazole preventive therapy; regimen 1: 2HRZE/4HR, regimen 2: 2SHRZE/1HRZE/3HRE (1, 2,3, 4: months, R: rifampicin, H: isoniazid, Z: pyrazinamide, Ethambutol; and S: streptomycin); MTB: Mycobacterium TB; RIF: resistance to rifampicin; and Alternatives: clinical diagnosis, score charts, chest X-Ray, magnetic resonance imaging (MRI), Mantoux test, or tissue pathology; N: total number of participants in the study; n: frequency; %: percentage of frequency; OR: odds ratio; Asterisks (\*):statistically significant at $p \leq 0.05$ ; CI: confidence interval

TB/HIV co-infected female patients had approximately 38% lower odds of an unsuccessful treatment outcome compared to male patients (COR: 0.62, 95%CI: 0.38-1.03). After adjusting for potential confounding variables, the odds of unsuccessful treatment outcome was still lower for female TB/HIV co-infected patients, but to a lesser extent (about 29%) compared to male patients (AOR = 0.71, 95%CI: 0.40-1.27). Patients aged 15-24 years, 25-34 years, and 45-55 years had significantly higher odds of unsuccessful outcomes, being 6.35 (95%CI: 1.56-25.86), 3.97 (95%CI: 1.37-11.45) and 6.40 (95%CI: 2.27-18.06) times more likely than children, respectively. Whereas there was no significant association between unsuccessful treatment outcome and the other age groups specifically, adults who fell within the age bracket of 35-44 years (AOR = 2.52, 95%CI: 0.98-6.48) and ≥55 years old (AOR = 3.65, 95%CI: 0.90-14.80), compared to the reference group.

TB/HIV co-infected patients who received ART had significantly 92% lower odds of an unsuccessful treatment outcome compared to those who did not receive ART (COR = 0.08, 95%CI: 0.01-0.67), but after adjusting for potential confounding variables, the odds of an unsuccessful outcome was still lower (about 89%) for TB/HIV co-infected patients who received ART (AOR = 0.11, 95%CI: 0.01-1.51). Patients who received CPT had significantly 79% lower odds of an unsuccessful outcome (COR = 0.21, 95%CI: 0.06-0.77). However, after adjusting for potential cofounders, those who received CPT had 21% lower odds of an unsuccessful treatment outcome (AOR = 0.79, 95%CI: 0.13-4.82), but this effect was not as strong as initially suggested by the COR.

Diagnosis through the alternative way was significantly associated with lower odds of unsuccessful treatment outcomes compared to smear microscopy (AOR = 0.32, 95%CI: 0.12-0.76). In contrast, Xpert MTB/RIF showed no significant association with treatment outcomes (AOR = 1.65, 95%CI: 0.64-4.27).

## Discussions

This study investigated patients with TB/HIV co-infection who received care at UPTH, Port Harcourt, Nigeria, providing insights into operational realities among the patients. The prevalence of HIV infection among registered TB patients in this study was 37.4%, which is much higher than the rates reported in other Nigerian tertiary hospitals, including 9.25% in Plateau [15], 21.6% in Lagos [18], 29.0% in Enugu [14], 29.3% in Ogun [32] and 33.9% in Imo [19]. The prevalence observed was also higher than rates reported in Ethiopia (17.7%) [33] and Ghana (18.0%) [34]. UPTH, being a referral hospital in the region, may contribute to the high TB/HIV co-infection rate, as it may attract patients with more complex cases. Additionally, the high proportion of HIV cases in Rivers State, where the UPTH is located, could also play a role, with the state ranking third among states with the highest HIV prevalence and accounted for 3.8% of HIV cases in Nigeria in 2019, according to the National Agency for the Control of AIDS [35]. The WHO’s recommendation to treat all patients with *Mycobacterium tuberculosis*, including those co-infected with HIV, may have also contributed to the high prevalence rate.

This study observed a treatment success rate of 52.2%, falling short of the national target of 85% set by the WHO [36]. In comparison, similar studies in Benue State [20] and Plateau State [15] reported higher treatment success rates of 68.9% and 87.5%, respectively. Whereas studies in Imo State and Abuja, however, reported lower treatment success rate of 41.1% [19] and 48.8% [17], respectively. The lost to follow-up rate among TB/HIV co-infected patients was 14.8%, suggesting that inadequate follow-up care and support may be a significant factor in reduced treatment success. This highlights the need for improved follow-up care and support for TB/HIV co-infected patients to achieve better treatment outcomes

Furthermore, the study observed an overall improvement in treatment outcomes for TB/HIV co-infected patients from 2014 to 2018, and a decline in treatment success rate in 2019. The odds of unsuccessful treatment outcome in 2019 was 31% lower compared to 2014, despite the 1.35 times higher COR. This shift underscores the importance of controlling for confounding factors to accurately assess temporal trends in treatment outcomes. Therefore, there is need for further investigation into the decline in treatment success rate in 2019 to deeply understand the underlying causes and implement corrective measures.

The mean age of TB/HIV co-infected patients in this study was 29.2 ± 18.9 years, which is lower than the mean age reported in similar studies from Nigeria [14-16,37,38], Tanzania [39], and Ghana [34]. The majority of TB/HIV co-infected patients in this study were below 15 years old (29.8%), while the least fell within the age bracket of 15 to 24 years (5.2%). Our findings is consistent with the study conducted in Lagos, Nigeria, which found that almost 30% of children under 15 years old were co-infected with TB/HIV [40]. It’s further supported by another study in Volta region of Ghana [41,42], but differs from studies in Ethiopia [43], and Ghana [44], which reported the higher prevalence of TB/HIV co-infection among adults aged 25-34 years. This age distribution differs from other studies, which reported the highest prevalence of TB/HIV co-infection among adults aged 30-39 years [14,39,45], 30-44 years [38], and 40-49 years [34]. Our study aligns with those of Zenbaba et al. [46], which reported the lowest TB/HIV co-infection rates among adults aged 15-24 in two public hospitals in Southeast Ethiopia. The high burden of TB/HIV in children under 15 years old compared to adults above 15 years, particularly those in the sexually active age groups of 25-34 years and 35-44 years, may be attributed to several factors. Mother-to-child transmission of HIV during pregnancy, childbirth, or breastfeeding is a significant contributor to the high burden of TB/HIV in children [47,48]. According to WHO [49], childhood HIV infection is particularly common in settings where antenatal HIV prevalence is high and prevention of mother-to-child transmission of HIV interventions are not widely implemented. These factors have been well-documented in previous studies, which have underscored their implications for TB/HIV transmission and diagnosis in children [48,50,51]. While these factors were not specifically examined in our study, they provide a plausible explanation for the observed high burden of TB/HIV in children under 15 years old.

Results in the multivariable logistic regression analysis indicate that age was statistically a significant predictor of treatment outcomes among TB/HIV co-infected patients. Remarkably, the 15-24 years, 25-34 years, and 45-54 years age categories were more likely to experience significant unsuccessful treatment outcomes, when compared to children under 15 years old. While the current study doesn’t provide direct evidence for the reasons behind unsuccessful treatment outcomes among these age groups, possible explanations based on existing literatures include differences in comorbidities, treatment adherence, drugs interactions or overlapping side effects of medications [52, 53].

Globally, the prevalence of HIV is typically higher among females compared to males [54]. However, our study found an almost equal prevalence of TB/HIV co-infection in both males (50.2%) and females (49.8%). This observation was in line with a previous study conducted at the National Hospital Abuja, Nigeria, which reported similar prevalence rates in male (42.7%) and female (42.6%) TB/HIV co-infected patients [17]. The observed pattern of equal prevalence in both males and females may be attributed to a shift in risky behaviors among men, potentially contributing to an increased risk of TB/HIV co-infection. This finding further shows the need for targeted interventions to address the changing dynamics of TB/HIV co-infection among demographic distributions.

This is a tertiary care which is reflected in many cases (about 41%) diagnosed in alternative ways. Our investigations revealed that patients diagnosed through these alternative ways had significantly better treatment outcomes, with 68% lower odds of unsuccessful outcomes compared to those diagnosed via smear microscopy. This indicates that alternative diagnostic approaches may facilitate earlier detection, particularly among TB/HIV co-infected patients. This early diagnosis likely enables clinicians to initiate timely treatment, thereby improving patient outcomes. Furthermore, these observations suggest that alternative diagnostic methods may offer advantages over traditional smear microscopy, potentially leading to enhanced patient care and treatment success.

Our study found that a high proportion of TB/HIV co-infected patients received ART (95.1%) and CPT (93.4%), which is consistent with findings from Kenya [55], but higher than studies conducted in Botswana [56] and Lagos, Nigeria [40]. The WHO recommends the use of ART and CPT, either singly or in combination, for TB/HIV patients [57]. The combination of ART and CPT has been shown to provide synergistic effects, reducing mortality and improving treatment success in TB/HIV co-infected patients [58-60]. Our study confirmed the importance of ART and CPT in enhancing treatment outcomes, with patients who received CPT more likely to have successful outcomes. Moreover, patients who had received either of the therapy exhibited reduced odds of unsuccessful treatment outcomes, suggesting that while ART or CPT was a crucial component of treatment approaches for TB/HIV co-infected patients, its impact may be modified by various individual or clinical factors. These findings have important implications for clinical practice and policy. They highlight the need for tailored treatment approaches that take into account individual patient characteristics and clinical contexts. By optimizing the use of ART and CPT, healthcare providers can improve treatment outcomes and reduce morbidity and mortality among TB/HIV co-infected patients.

### Limitation of the study

This study had several limitations inherent to its retrospective design. We relied on existing patients’ medical records from a federal tertiary-care facility in Port Harcourt, which may be prone to underreporting, missing data, and errors. To mitigate this, we independently implemented double data entry and checked for inconsistencies. However, the registers did not contain information on reasons for loss to follow-up, transferred out or mortality rates. Furthermore, our analysis was limited by the unavailability of data on potential confounding factors, such as educational level, patient lifestyle, socioeconomic status, comorbidities, treatment adherence, severity of immune suppression, timing of ART and CPT, drug adverse effects and some other relevant variables. This precluded us from adjusting for competing risks and drawing causal inferences regarding the impact of these factors on treatment outcomes. While findings may not generalize to other states or regions in Nigeria, our sample’s representativeness of TB/HIV co-infection prevalence in Port Harcourt reduces concerns about systematic differences from the broader populations in Nigeria. Future prospective studies building on these findings could further elucidate the dynamics of TB/HIV co-infection, informing evidence-based practices and policy decisions.

## Conclusions

The prevalence of TB/HIV co-infection is high in the study population, exceeding rates reported in some other Nigerian tertiary-care hospitals. The treatment success rate of 52.2% falls short of the national target of 85%, emphasizing the need for improved follow-up care and support. Notably, children ≤14 years old bear a significant burden of TB/HIV co-infection, likely due to mother-to-child transmission. Public health strategies and targeted interventions are crucial to addressing the changing dynamics of TB/HIV co-infection, particularly among high-risk age groups. Study demonstrates the benefits of alternative diagnostic methods, ART, and CPT in enhancing treatment outcomes among TB/HIV co-infected patients. The integration of these interventions into clinical practice and policy has the potential to reduce mortality rates and improve treatment success.

## Data Availability

All relevant data are within the manuscript and its Supporting Information files.

## Funding sources

This research did not receive any specific grant from funding agencies in the public, commercial, or not-for-profit sectors.

## Acknowledgements

The authors wish to thank Center for International Health (CIH), University of Bergen, Norway, and Public Health Society Against Infectious Diseases (PHSAID), Nigeria for their support and guidance throughout this study. We also thank all staff and management of the Department of Community Medicine, UPTH, especially those in the DOTS clinic who gave us access to the register.

## Author contributions

PCN: Conceptualization; Data curation; Formal analysis; Investigation; Methodology; Project administration; Validation; Visualization; Writing - original draft; Writing - review editing. SCN: Data curation; Investigation; Writing - review editing. ATA: Data curation; Writing - review editing. INO: Data curation; Writing - review editing.

## Notes

### Competing Interest Statement

The authors have declared no competing interest.

### Funding Statement

The author(s) received no specific funding for this work.

### Author Declarations

UPTH Ethics and Research Committee (ERC), Nigeria, and Regional Committee for Medical and Health Research Ethics (REK), Norway.

